# Investigating a patient-led conference: What are the characteristics and impacts of patient leadership? A qualitative study

**DOI:** 10.1101/2025.11.07.25339430

**Authors:** Tianna Magel, Kimberly Strain, Anna Samson, Dawn P. Richards, Hetty Mulhall, Karim Khan, Lorelei Lingard

## Abstract

**Background:** Patient engagement has been implemented in various settings including clinical, research, and quality improvement, with varying levels of patient contributions and decision-making responsibility. However, little is known about the experiences of patient partners who are in leadership roles in patient-led events. For Patients, By Patients (PxP) is an annual, virtual, patient-led conference that focuses on topics important to patient partners in research. Each year’s PxP steering committee is comprised of those with patient experiences and consequently, offers an opportunity for our research team to explore patient leadership within a conference setting. Understanding more about the intricacies of patient-led events is necessary if we wish to support patient leadership as a valuable form of patient engagement.

**Objectives:** The aim of this study was to address the current knowledge gap in patient-led events and patient leadership.

**Design:** We conducted a qualitative descriptive study of semi-structured virtual interviews with PxP conference steering committee members. Thematic analysis was used to identify core themes that were salient to the data.

**Setting:** International virtual setting via Zoom from Jan 2025-April 2025.

**Participants:** Purposive sampling was used to conduct interviews with thirteen PxP patient partner steering committee members.

**Results:** Four core themes were identified in the data: institutional support, steering committee environmental characteristics, personal growth, and new possibilities.

**Conclusions:** Patient-led events offer an opportunity to promote patient leadership. To facilitate patient leadership in this setting, several factors are important including attention to power dynamics, institutional support, and considerations for accessibility and intersectionality.

## Introduction

Patient engagement has been increasingly recognized and valued as a core component of quality research.^1–3^ There have been calls from research communities and patient partners to have patients hold leadership positions within healthcare, research teams, and grants.^4–8^ However, patient leadership is still an emerging area and there are very few published examples of best practice for patients leading and holding power in the research ecosystem.

Patient leadership aims to build on the patient engagement spectrum to promote patient partner autonomy and ability to influence or direct decision-making.^6,9–11^ Patient leadership is also referred to as consumer leadership in Australia or patient and carer leaders in the UK.^5,12^ Those in leadership roles are “recognized as service leaders, equal in esteem and influence to managerial and clinical leaders” where they are treated as equals and not tokenized.^12^

Despite calls for patient leadership from patient partners and members of the research community, the practice of patient leadership is not well characterized or defined. With varying opinions on how to best implement patient leadership,^13^ clarity of concept is essential. However, patient leadership is poorly represented in the literature, which tends to focus on more “traditional” engagement roles, such as consultation and co-creation.^9^ While there is knowledge of patient leadership experiences held within the patient partner community, this evidence base continues to be poorly accepted by others in the academic space.^14–16^ Outside of previous literature detailing our own event, we found no literature on patient-led conferences about patient engagement in research.^14–17^ Furthermore, in other disease-specific events with patient leaders, the speakers were almost exclusively health professionals—patient conference leaders seemed to choose the speakers (often health care practitioners) but did not appear to have a clear speaking role in the program.^15,16^

The annual international, virtual, “For Patients, By Patients” (PxP) conference, that began in 2023, is a patient-led conference. Rather than focusing on any particular disease(s), PxP focuses on patient engagement in research. The PxP conference infrastructure and funding is supported by the Canadian Institutes of Health Research (CIHR) - Institute of Musculoskeletal Health and Arthritis (further referred to as the Institute). Information on the structure of CIHR and its thirteen institutes can be found elsewhere.^18^ To ensure patient partners had the autonomy to make decisions, they were intentionally kept at arms-length from the Institute. With support from the patient partner facilitator, the content and speakers for the conference were decided upon by the patient partner steering committee (i.e., decisions were entirely patient led). A horizontal leadership approach (i.e., where leadership roles are shared) was used to ensure steering committee members could come to decisions independently and collaboratively, with minimal input from the Institute.^19,20^ Further details on how the PxP conference was established and run are described elsewhere.^17^

Because there is no literature on issues such as the challenges that patient leaders have in their roles in research, how they resolve conflict, whether they perceive power imbalances, we endeavored to focus on the experiences of patient partner steering committee (SC) members from two cohorts of the PxP conference. Our aim was to better understand: (i) patient partner perspectives of this patient-led event and (ii) how to better support patient leadership.

## Methods

Our study followed a descriptive qualitative research design, appropriate for underexplored phenomena and work that prioritizes detailed description of experience over theory-building or abstraction.^21^ Participants in this study were purposefully sampled on the basis of their experience being on the PxP conference steering committee, following information-power sampling approaches.^22^

Data collection and analysis occurred simultaneously, with previous interviews and preliminary insights used to inform future data collection. Thematic analysis, a flexible analytical tool that is compatible with descriptive qualitative research, was used to identify recurring themes in the transcripts.^23^ Data collection was considered complete once the research team reached a level of thematic sufficiency as part of the ongoing data analysis.^24^ Thematic sufficiency was achieved when patterns within the data were judged to be consistent and well-elaborated, and no new themes were identified as part of further analysis.

### Participants

We conducted purposive sampling to explore the leadership experiences of patient partner SC members from North America, Europe, Africa, and Oceania who were involved in the international annual PxP patient-led conference. All SC members (n=17) from the 2023 (n=9) and 2024 (n=8) PxP Conferences were invited via email by the Study Lead (TM) to participate in the study. Of the 17 members, 13 gave written consent and were recruited into the study, two expressed interest but subsequently did not respond, and two did not respond to invitations to participate.

SC members joined the PxP steering committee with varying degrees of prior patient engagement experience (e.g., number of past patient experiences in healthcare and research, living in places where patient engagement is new vs established, and whether those patient engagement experiences were positive or negative). These differing degrees of experience were captured in our sample. Prior experiences often served as an explicit point of comparison for SC members, and may also have served as a subconscious point of reflection, when discussing their steering committee experiences, what the PxP had done well, as well as areas for improvement. Because PxP was patient-led, the SC members (i.e., patient leaders) were well equipped to comment on their experiences and provide novel insights into patient leadership in research.

### Study setting and data collection

This study was conducted between January 2025 to April 2025. A semi-structured, open-ended interview guide was co-developed with patient researcher partners, KS and AS (see supplemental file 1). In-depth virtual interviews with SC members were conducted and co-led by TM with either KS or AS. Interviews were 1-hour in duration and took place virtually on Zoom. For their participation, SC members were offered an honorarium in accordance with CIHR-IMHA Patient Partner Compensation Guidelines.^25^

### Data analysis

All interviews were recorded and the audio automatically transcribed verbatim via Zoom. TM reviewed and manually edited all interview transcripts to ensure accuracy. Field notes from both co-interviewers were written and used to inform analysis. Interview transcripts were first reviewed by TM, KS, and AS to establish familiarity with their content, followed by an initial round of coding conducted by TM, KS, and AS. In line with Braun and Clark’s 6-step process, initial codes were established, reviewed, and refined through an iterative process with the entire study team.^23^ Codes were grouped into emerging themes defined by the study team over multiple meetings. Coding development and theme refinement took place until no new codes were generated. All analysis of the data was conducted via NVivo V.14. We have also followed the Consolidated Criteria for Reporting Qualitative research guidelines to increase the transparency of our findings (see supplemental file 2).

## Positionality

In qualitative research, the research team must attend to how its orientations shape the study and influence the credibility of the findings. Our research team included both individuals intimately involved in the patient engagement space and the annual PxP conference. AS is a patient partner and is a current member of the 2025 PxP steering committee. KS is a patient partner and was a panel member for a session at the PxP conference as well as moderator for a panel discussion during a PxP webinar. DPR identifies as a patient partner, PE specialist and facilitator, facilitating PxP steering committee meetings since 2023, and provided support to speakers, moderators and SC members in their roles at PxP. The lived experiences of AS, KS, and DPR as patient partners brought important insights and nuance to the interpretation of the data. HM attended steering committee meetings to engage with the process and provide logistical support. She managed the PxP website, social media and event platform, and also supported speakers during the event. She was intimately involved in developing PxP and steering committee meetings. TM attended several PxP conferences and webinar series. TM has also worked closely with patient partners on various Institute projects and is a supporter of patient engagement in research. KK is an experienced clinician-scientist who has been part of many events as a funder, organizer and participant. He is an advocate of patient engagement in research. He attended some of the PxP planning meetings and met regularly with DPR and HM. The team also included LL, a qualitative methodologist uninvolved with PxP, who provided guidance for the study design and participated in the iterative cycles of data analysis. The prior patient engagement experiences, research expertise, and collaborative nature of this study team allowed for in-depth discussions as analysis proceeded and enriched the interpretation of the results.

## Patient and Public Involvement in Research

Patients and the public were Co-Investigators. Patient Research Partners co-designed all aspects of the study from the beginning, including developing the research questions, co-leading the interviews with SC members and data analysis, and co-authoring this manuscript. Patient Research Partners were invited to participate in all team meetings and were compensated for their time in accordance with CIHR-IMHA compensation guidelines.^25^

## Results

Descriptive thematic analysis identified four major themes within the data: (i) Institutional Support, (ii) Steering Committee Environmental Characteristics, (iii) Personal Growth, and (iv) New Possibilities (see Figure 1). Each theme is described below and illustrated with representative quotes from the transcripts.

**Figure 1.**
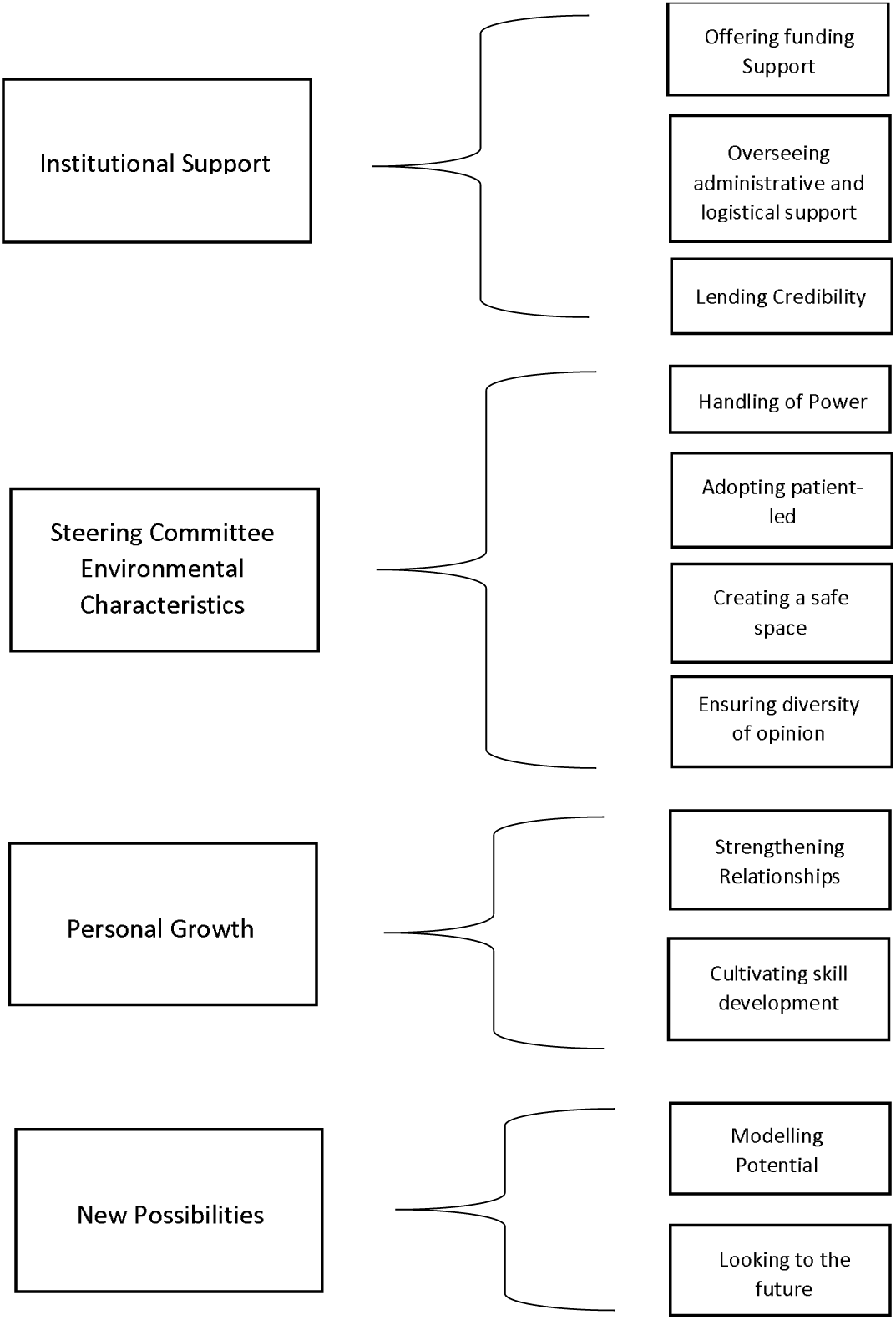
Patient-led Themes.

### Theme 1: Institutional Support

SC members identified institutional support as an essential resource to support the steering committee members and to make the PxP conference happen. This included offering financial compensation to SC members for their time and expertise, which SC members felt reflected the importance of patient engagement to the Institute, that participant time was valued and their input “actually meant something” (SC member 4).

> *A lot of people say that patient partnerships or patient-led work or whatever are really important to them, but they don’t ever put their money where their mouth is. (SC member 1)*

SC members also highlighted that the institutional support of PxP provided a level of credibility when interacting within the research community. Often there are barriers to patient partner events being taken seriously by the research community when “someone does not have institutional backing” (SC member 3). Buy-in from a reputable institution lent legitimacy to the event and the information shared by speakers, many of whom were patient partners.

> *Sometimes, particularly in terms of researchers joining the event, I think if it was organized by a patient charity or some patient group, I don’t think it would have that same gravitas that researchers think. ‘Oh, this is something I want to join or this is something for me’ … But I think if it’s got a big institution behind it. I think that helps with broadening the appeal. (SC member 13)*

SC members described the administrative and logistical support as “exemplary” (SC member 9), differing positively from their other patient engagement experiences. The additional administrative support also offered patient partners an overall sense of smoothness with the experience. SC members and conference speakers were able to schedule one-on-one or additional meetings with the patient partner facilitator. These meetings accommodated SC members in different time zones and provided opportunities to ask questions and share things they might not have wanted to in group discussions. Overall, SC members felt a sense of trust in the administrative support and this reduced the burden on patient partners allowing them to show up to meetings without feeling stressed or inconvenienced.

> *It’s basically… the dream support we had. There was process support, there was content support, there was tech support. I mean, everything was sorted and everything was handled. (SC member 2)*

### Theme 2: *Steering Committee* Environmental Characteristics

SC members identified several characteristics of the PxP steering committee environment that were salient to the committee and event success. Of these, being patient-led was highlighted as a core characteristic by the steering committee. In their own words, SC members defined what they felt were the “key ingredients” to ensuring events or projects were truly patient-led. These included patient partners: driving the initiative and defining the direction; originating the ideas for research or projects; being embraced as equals and having a shared partnership on the team (i.e., having a “seat at the table” (SC member 9)); and being involved in the implementation of a project from start to finish. SC members highlighted that patient-led events should be just that, primarily led by patients.

> *So, when I think of patient-led or something that is patient co-led. It’s not just about participating in research. It’s not just about partnering from being an advisory committee or being on a working group. It’s not just advocating for policy changes or legislative changes or a new drug or voice in something. It’s about being an equal leader and having strategy, having blue sky thinking, defining direction, moving forward with implementation, making choices, making decisions in a democratic way. (SC member 11)*

Some SC members had never seen or been a part of leading an entirely patient-led event outside of PxP. SC members stated they felt they were able to have a say in various aspects of the conference and that the experience was so positive that it left them wondering when the “other shoe would drop” (SC member 2). Overall, the majority of SC members felt that the steering committee and decisions that emerged from the group were patient-led.

> *The reason it’s different is that it is different. It’s genuinely patient-led. (SC member 2)*

Various themes on how “power” was handled were woven into the reflections of SC members about their steering committee experiences. Conversations around holding and sharing power mainly fell into one of the following categories: decision-making power (i.e., who holds the power to make decisions), power conventions (i.e., power systems that develop within groups when no clear hierarchy exists), and managing power (i.e., institutions giving or sharing power with patient partners). SC members expressed that their decision-making capacity was reinforced by leadership “sharing” (SC member 6) or “giving up” (SC member 1) power. Those in positions of power within the Institute were noted as providing financial and moral support but having a hands-off approach that did not control the narrative of the steering committee nor its decisions.

> *They were so supportive. But they didn’t control it. They let us drive the bus, so to speak. (SC member 5)*

Several subthemes around power conventions emerged, reflecting how power systems can develop within groups when there is no clear leader or final decision maker. Institute staff aimed to equalize power and establish horizontal leadership among SC members and Institute leadership. However, several members indicated that their ability to express their opinions or have their input considered by other SC members was challenging at times based on the dynamics of the group. Indeed, some SC members felt their voice was lost or diluted during the group discussion, while others felt intimidated in group discussions and expressed challenges in initially forging connections over virtual meetings.

> *But here, because everyone was a patient, I sometimes felt like people were fighting to be heard above the next person. We’re all on the same level field so then people want to kind of stand out a bit sometimes. (SC member 13)*

In contrast to this, the absence of a hierarchy simultaneously cultivated a space that was collaborative, open, and respectful. SC members expressed that the steering committee was a positive and open environment where opinions were heard and no topic was off limits. SC members were not “afraid” of being constrained or “shot down” (SC member 3) by institutional leadership. SC members emphasized the impact leadership from the Institute and the Patient Partner Facilitator had in fostering trust and respect within the steering committee. This allowed SC members to feel a sense of openness and that their contributions were acknowledged without being micromanaged.

> *It makes a huge difference when the leader sets the tone … of “this is your project”. (SC member 1)*

The diversity of disease expertise and global representation of steering committee members was also acknowledged as a vital component to ensuring representation and accessibility. SC members identified accessibility and global representation as essential to the conference having a bigger impact by reaching beyond the “usual suspects” (SC member 2) and having discussions around a greater variety of topics with more international reach. SC members reflected on prior patient engagement experiences where they felt their voices and opinions were not heard, often describing feelings of being tokenized when engaging with the medical community or researchers.

> *I always take that away from PXP. I got to see exactly what a diverse cohort looks like. So, when folks talk about health equity. I can go a little deeper because you’re talking about it within the confines of [location]. But I actually had the opportunity to participate on real diversity, equity, and inclusion and it wasn’t about race. (SC member 9)*

However, some SC members emphasized the need for the inclusion of perspectives outside of the Global North and for the consideration of cultural differences as well as the inclusion of those from different educational backgrounds.

Flexibility and consideration for accommodating steering committee needs and being able to pitch in according to each member’s strength were highlighted as fundamental to meeting accessibility needs. Rescheduling or offering additional meetings, materials in advance, and accessible features of the conference, allowed SC members to feel like they were not missing out and were included and valued in the experience.

> *The expectations were very clearly communicated that sort of no one had to do more than they felt able to. (SC member 2)*

### Theme 3: Personal Growth

SC members highlighted that the various methods of engagement in the steering committee helped foster personal growth and the development of practical and hands-on skills. Often SC members reflected on the positive experience they had with the PxP steering committee and described how their involvement gave them “a push to do more” (SC member 10) and fostered a sense of confidence in their expertise and ability to “lead other initiatives” (SC member 7). SC members reflected on how the hands-on skills they developed from the steering committee led to feeling more confident in using their voice to share their opinion, get involved in conferences, and advocate for their own condition. They also highlighted how this experience provided an opportunity for reflection on what they could do in their own country or community.

> *I gained confidence and made connections and even presenting, I gained confidence in my presentation skills and my facilitator skills. (SC member 5)*

The majority of SC members also expressed that involvement with the steering committee fostered peer-to-peer support, relationship building, and “a kinship” (SC member 9) with other members of the committee and patient partner community. Many felt that they were able to connect and build a sense of community and “partnerships that continued” (SC member 11) past the conclusion of the PxP conference. Several SC members shared that their involvement with the conference also made them feel more connected to the health research community and, in several instances, resulted in researchers who were attending the conference reaching out to collaborate on future projects.

> *I would say it’s made me feel more present within the health research community… it’s made me feel more included in health research. It feels like more opportunities have opened up for me as a patient partner since then. (SC member 3)*

### Theme 4: New Possibilities

Several SC members noted that the Institute is an innovator in the patient engagement space and is leading by example. This impact was perceived by SC members as having a “ripple effect” (SC members 2 and 4) and showing other institutions what is possible. SC members noted there are very few patient-led events. Bringing individuals together from around the world encourages patient partners and others to go back to their organizations or countries and advocate for patient engagement. SC members also spoke about how patient-led events can improve future research by increasing awareness and educating other researchers about patient engagement best practices.

> *PxP is really showing the industry that… you for so many years, you’ve forgotten us. And we had to create a forum. To let you know that we can do this … Our purpose is to educate you and speak for those that aren’t here on what some of the disparities and some of the challenges are when it comes to helping us to live better, healthy lives. (SC member 9)*

SC members encouraged future steering committee members to have confidence and ensure their ideas are heard. They also encouraged the Institute to keep forging ahead with patient-led events and to keep expanding. SC members also stressed that “[patient engagement] happens by choice rather than chance” (SC member 6), highlighting the need for the same level of promotion of patient engagement and patient-led events across all of CIHR’s Institutes. Some SC members voiced concerns that the event wasn’t reaching enough individuals and expressed a need for greater promotion of the PxP conference to engage a larger audience.

> *I hope they continue something like this … [the PxP conference] can really benefit a lot of people. To bringing the patient autonomy voice forward… allowing them to have the autonomy to choose … how and what information is presented from the heart and the mind. (SC member 7)*

## Discussion

The goal of our study was to better understand how patient partners in leadership roles experienced a patient-led conference and to start characterizing best practices for institutes and others to support patient partners in leadership roles. Interviews with SC members highlighted four major themes that were vital to the success of the patient-led PxP conference and patient partner leadership experience: (i) institutional support, (ii) steering committee environmental characteristics, (iii) personal growth, and (iv) new possibilities. Here we aim to expand on the first two themes, and to discuss important insights relating to institutional support, power dynamics, accessibility and intersectionality.

### Providing institutional support

Events such as PxP have costs and require personnel support—this can be provided by an organization or institution. Without these supports from the Institute, it would have been very challenging to run a patient-led event of the same caliber.^17^ Until patient partners are given the necessary support and funding to run their own events, successful patient leadership will rely on a level of support from institutions. In addition, when hosting or leading an event, patient partners face unique barriers related to their health and life circumstances, as compared to typical conference organizers. Our results suggest that these barriers can be navigated through additional and flexible logistical supports and meeting times, allowing patient partners the opportunity to focus their time and energy on important aspects of the event, such as the content and speakers, rather than worrying about the complexities of technology and scheduling etc.

### Patient leadership

While a patient-led event may require financial support, the leadership remains with patient partners. Financial support without strings is essential; this can be realized through a defined budget (and potentially in-kind support such as staff) and a clear mandate that patient partners are in charge. Our results suggest that patient leadership need not be limited only to patient-led conferences; rather, it should be encouraged in areas including research design, health policy, advocacy for access to care, and clinical care design.

The level of support provided can exist on a continuum. There are several examples of patient-led initiatives that range in their level of organizational involvement and support, including Pain Canada’s “Putting the Pieces Together” conference, ^26^ which served as a catalyst and template for the PxP conference, and the “Canadian Arthritis Summer School”.^27^ Both of these events supported patient partners to lead various aspects of decision-making and the event proceedings.

There are also examples of patient leaders spearheading their own initiatives without institutional support, such as the Australian Consumer Partnerships in Research Awards.^28^ Additionally, in 2025, the Canadian-based PxP expanded to include PxP Africa, India, and Australia. These events have operational and financial support from CIHR-IMHA so that SC members in those regions can create, lead, and host an event that meets the needs of their region. Each of these events is supported without any expectations beyond “create an event for your region about patient engagement in research.” Ultimately, our results suggest that providing monetary, logistical and personnel support is a way for organizations to demonstrate that they value patient leadership but also, that they trust patient partners and that they can provide resources, without any strings attached, and know that patient partners hold the skills and abilities to take charge and lead events.

### Power and Power Dynamics

Power among committee members was a central component discussed by SC members in this study, and a complex one. Efforts to eliminate hierarchies had the unexpected impact of complicating the decision-making process. Richards et al., previously described power and power dynamics in patient engagement, including the changing nature of power, power amongst and between the patient partners themselves, tokenism, and patient engagement being viewed as a commodity.^29^ These assertions about power resonate with SC member reflections on prior patient engagement experiences. However, efforts to create an equal and collaborative space between the steering committee and leadership raised the issue of what happens when you alter traditional power structures? In these situations, “traditional” decision-making rules no longer applied and exposed some unintended consequences.^30^ Often in healthcare, and sometimes research, patient partners have the least amount of power, and disrupting this hierarchy revealed further power dynamics that need to be considered, particularly when examining the intersectionality that each SC member brings.^29^ This shift in power may have also pushed SC members to rely on what they know, and what is comfortable. As highlighted by Carolyn Thompson, a “sudden shift to a new order can shatter things people previously counted on as solid”.^31^ As a result, power structures within the SC may have formed out of familiarity and a need for some form of stability. It may take time, clear communication, and regular check-ins with SC members to ensure that each member feels equally comfortable asserting themselves in a decision-making process with unconventional power relations.

SC member reflections also highlighted the varying degrees to which individuals recognized their own power and positionality within the committee. Some SC members felt that their opinions were heard and integrated into actions, while others did not. This may have reflected differing positionalities within the group, not realizing that some members did not share the same experience.^32^ Many SC members are from parts of the world where patient engagement is better established and often had advanced education. Thus, even with the most intentional efforts to “level the playing field” the intersectionality of participant characteristics (e.g., education, wealth, language etc.) and differing degrees of privilege, may ultimately create an unintentional power dynamic within the SC. These reflections on power systems, and what happens when they do not follow the status quo, highlight the complicated nature of trying to democratize power and the subsequent roles individuals may try to fill when no named leader exists.

### Accessibility

Each year of the PxP SC has grown more global and has taken intentional steps to broaden diversity.^33^ As PxP continues to expand, future approaches to facilitating meetings may need adaptation in favor of accommodating SC members from diverse cultures and various parts of the world.^34,35^ In line with this, one characteristic of the environment that was particularly important was accessibility. Meeting virtually and adopting a flexible meeting schedule allowed SC members from around the world to contribute in ways that worked for them, removing geographical and time constraints. However, while virtual settings have been noted to increase accessibility,^36^ they also bring their own set of challenges. Individual learning curves navigating online platforms as well as precarious access to the internet can create barriers for international patient partners.^37,38^ These challenges should be discussed with patient partners, and accommodations, such as potential compensation for internet access (e.g., via mobile networks), should be considered.

### Take home messages

Below (see Table 1) we consider the above reflections and offer some pragmatic suggestions on key areas to be open about and discuss with patient partners.

**Table 1.** Institute lessons learned and recommendations.

| **Institute lessons learned** |  |  |
| --- | --- | --- |
| **Topic** | **Lesson Learned** | **Future Considerations** |
| 1. Let Patients Lead | Financial, leadership, and institutional support are needed to run the event and support patient leaders. | Ensure adequate support is provided in all aspects of the event, including patient partner compensation, logistical support, staffing, and technology. |
| 2. Democratize Tasks and Decisions | Power dynamics and power hierarchies can develop even when “traditional” rules and procedures are dismantled. All patient partners should have an equal say in the decision-making process. | Spend time on your decision-making process. Discuss with patient-leaders how traditional rules and procedures have been dismantled and what has been put in their place. |
| 3. Make the Event Accessible for Patient Partners | Accessibility encompasses a number of factors such as a patient partners’ language, education level, time zone, internet access, and ability to be present at meetings and engage synchronously vs. asynchronously. | Discuss and accommodate patient leaders’ accessibility needs and preferences for meeting frequency, time, and setting (i.e., group vs. 1:1). |
| 4. Acknowledge Intersectionality | Consider Intersectionality when engaging patient leaders from different places and backgrounds. | Ask patient leaders about their communication style and preferences. Ask patient leaders how you can support them to lead engagement with their own community. |

## Limitations and Future Directions

Our sample size is small; however, it is representative of the population we aimed to study and information rich. In line with the concept of information power, our sample size makes up most of the population of interest (n = 13/17), the quality of dialogue was strong, and our sample specificity was dense.^22^ Additionally, our analysis was iterative and informed by our team which included Patient Research Partners. Our findings are limited to the learnings from two years of the same annual patient-led conference. Furthermore, SC members were mostly from places where patient engagement is already fairly well-established. Because two years have passed since some SC members initiated their role, their reflections may be deeply impacted by their own patient partner experiences since that time, current life circumstances, and health. However, given the novelty of this research area and the limited number of international patient-led events, these findings offer a valuable opportunity to add to the current literature on patient leadership and patient-led conferences. Future research should expand beyond conferences and examine patient leadership in different contexts, including healthcare, policy, and advocacy settings.

## Conclusion

In our descriptive thematic analysis, we found several components that supported patient leadership in a patient-led conference. Offering appropriate support, including financial compensation, creating a safe, accessible, and culturally diverse space, as well as providing opportunities for skill development and relationship building were all important aspects of patient leadership. Patient-led events also offer an opportunity for institutes and the wider researcher community to place their trust in patient partners, allowing them the autonomy to make decisions that are important to them and their community. We offer several lessons learned and recommendations with the hope that our findings encourage institutes and researchers to recognize the value patient leadership roles can add and to propel patient-led initiatives to continue expanding to other events, policy, and research. Overall, our findings underscore that patient partners can lead a successful virtual conference.

## Supporting information

Supplemental File 1. Interview Guide

Supplemental File 2. COREQ Checklist

## Data Availability

Our research ethics board approval requires that transcripts be viewed only by the research team. The data are not publicly available to protect the privacy of study participants.

## Author contributions

All authors met the International Committee of Medical Journal Editors (ICMJE) criteria for authorship.

All authors contributed to the conception, design, and interpretation of the work. TM, KS, and AS performed data collection and analysis. All authors made substantial contributions to drafting and revising the work.

## Competing Interests Statement

AS is a steering committee member for the PxP 2025 conference and received an honorarium in line with CIHR-IMHA guidelines for their involvement on the research team.

DPR is the facilitator for PxP steering committee meetings and provides logistical support.

HM attended steering committee meetings to provide logistical support and managed the PxP website and social media.

KK was the Scientific Director of the CIHR Institute of Musculoskeletal Health and Arthritis from 2017-2025 (e.g. during the period that this research was undertaken) and attended steering committee meetings to provide logistical support.

KS received honoraria in line with CIHR-IMHA compensation guidelines for her involvement on the research team, as well as for her involvement in the 2024 PxP Conference and webinar.

TM attended several PxP 2024 conference sessions and was an employee of CIHR’s Institute of Musculoskeletal Health and Arthritis at the time of this research.

LL has no competing interests to declare.

## Ethics Statement

Ethical Approval was obtained from the University of British Columbia Behavioural Research Ethics Board (H24-02930).

## Funding statement

This work was supported by Canadian Institutes of Health Research Operating Grant SOP-154942

## Acknowledgements

We would like express our deepest gratitude to the 2024 and 2025 PxP Steering Committee members who participated in this study for their invaluable insight and contributions.

## Notes

### Author Declarations

The Behavioural Research Ethics Board of the University of British Columbia gave ethical approval for this work.

## References

1. Domecq JP, Prutsky G, Elraiyah T, et al. Patient engagement in research: A systematic review. BMC Health Serv Res. 2014;14(1):1–9. doi:10.1186/1472-6963-14-89/FIGURES/3

2. Forsythe LP, Carman KL, Szydlowski V, et al. Patient Engagement In Research: Early Findings From The Patient-Centered Outcomes Research Institute. Health Aff (Millwood). 2019;38(3):359–367. doi:10.1377/HLTHAFF.2018.05067

3. Vat LE, Finlay T, Jan Schuitmaker-Warnaar T, et al. Evaluating the “return on patient engagement initiatives” in medicines research and development: A literature review. Health Expectations. 2020;23(1):5–18. doi:10.1111/HEX.12951

4. Scholz B, Stewart S, Pamoso A, Gordon S, Happell B, Utomo B. The importance of going beyond consumer or patient involvement to lived experience leadership. Int J Ment Health Nurs. 2024;33(1):1–4. doi:10.1111/INM.13282

5. Scholz B. We have to set the bar higher: towards consumer leadership, beyond engagement or involvement. Australian health review. 2022;46(4):509–512. doi:10.1071/AH22022

6. Roper C, Grey F, Cadogan E. Co-Production: Putting Principles into Practice in Mental Health Contexts.; 2018.

7. ResearchNet. Catalyst Grant: Oral Health Data Analysis. 2024. Accessed September 18, 2025. https://www.researchnet-recherchenet.ca/rnr16/vwOpprtntyDtls.do?all=1&masterList=true&next=1&org=CIHR&prog=4240&resultCount=25&sort=program&type=EXACT&view=currentOpps&language=E

8. ResearchNet. Catalyst Grant: Digital Health. 2025. Accessed September 18, 2025. https://www.researchnet-recherchenet.ca/rnr16/vwOpprtntyDtls.do?all=1&masterList=true&org=CIHR&prog=4381&resultCount=25&sort=program&type=EXACT&view=currentOpps&language=E

9. Stewart S, Scholz B, Gordon S, Happell B. ‘It depends what you mean by leadership’: An analysis of stakeholder perspectives on consumer leadership. Int J Ment Health Nurs. 2019;28(1):339–350. doi:10.1111/INM.12542

10. Health Quality BC. IAP2 Spectrum of Public Participation.; 2024.

11. Carman KL, Dardess P, Maurer M, et al. Patient and family engagement: A framework for understanding the elements and developing interventions and policies. Health Aff. 2013;32(2):223–231. doi:10.1377/HLTHAFF.2012.1133/ASSET/IMAGES/LARGE/2012.1133FIGEX1.JPEG

12. Mcnally D, Sharples S, Craig G. Patient leadership: Taking patient experience to the next level? Patient Exp J. 2015;2(2):7–15. doi:10.35680/2372-0247.1091

13. Scholz B, Gordon S, Happell B. Consumers in mental health service leadership: A systematic review. Int J Ment Health Nurs. 2017;26(1):20–31. doi:10.1111/INM.12266

14. Rhodes P, Nocon A, Booth M, et al. A service users’ research advisory group from the perspectives of both service users and researchers. Health Soc Care Community. 2002;10(5):402–409. doi:10.1046/J.1365-2524.2002.00376.X

15. Levy M. Patient led conferences: an education for doctors. BMJ : British Medical Journal. 2001;323(7307):279. Accessed September 18, 2025. https://pmc.ncbi.nlm.nih.gov/articles/PMC1120889/

16. Zakrzewska JM, Jorns TP, Spatz A. Patient led conferences--who attends, are their expectations met and do they vary in three different countries? European journal of pain. 2009;13(5):486–491. doi:10.1016/J.EJPAIN.2008.06.001

17. Richards DP, Mulhall H, Belton J, et al. Co-creating and hosting PxP: a conference about patient engagement in research for and by patient partners. Res Involv Engagem. 2024;10(1):1–12. doi:10.1186/S40900-024-00603-0

18. Canadian Institutes of Health Research. Organizational structure. March 18, 2025. Accessed September 18, 2025. https://cihr-irsc.gc.ca/e/37926.html

19. Yu M, Vaagaasar AL, Müller R, Wang L, Zhu F. Empowerment: The key to horizontal leadership in projects. International Journal of Project Management. 2018;36(7):992–1006. doi:10.1016/J.IJPROMAN.2018.04.003

20. Crevani L, Lindgren M, Packendorff J. Leadership, not leaders: On the study of leadership as practices and interactions. Scandinavian Journal of Management. 2010;26(1):77–86. doi:10.1016/J.SCAMAN.2009.12.003

21. Sandelowski M. What’s in a name? Qualitative description revisited. Res Nurs Health. 2010;33(1):77–84. doi:10.1002/NUR.20362

22. Malterud K, Siersma VD, Guassora AD. Sample Size in Qualitative Interview Studies: Guided by Information Power. Qual Health Res. 2016;26(13):1753–1760. doi:10.1177/1049732315617444

23. Braun V, Clarke V. Using thematic analysis in psychology. Qual Res Psychol. 2006;3(2):77–101. doi:10.1191/1478088706QP063OA

24. Braun V, Clarke V. To saturate or not to saturate? Questioning data saturation as a useful concept for thematic analysis and sample-size rationales. Qual Res Sport Exerc Health. 2021;13(2):201–216. doi:10.1080/2159676X.2019.1704846

25. CIHR Institute of Musculoskeletal Health and Arthritis. Patient Compensation Guideline.; 2024.

26. Pain Canada. Putting the Pieces Together. 2025. Accessed September 18, 2025. https://www.paincanada.ca/events/putting-the-pieces-together

27. CIHR Institute of Musculoskeletal Health and Arthritis. Canadian Arthritis Summer School 2025. 2025. Accessed September 18, 2025. https://www.cihr-irsc.gc.ca/e/54046.html

28. AccessCR. Australian Consumer Partnerships in Research Awards 2025. August 2025. Accessed September 18, 2025. https://ccrew.accesscr.com.au/cpir-2025/

29. Richards DP, Bowden J, Gee P, et al. The ultimate power play in research - partnering with patients, partnering with power. Res Involv Engagem. 2025;11(1):1–9. doi:10.1186/S40900-025-00745-9/TABLES/1

30. Sweeney LB. Systems Thinking: A Means to Understanding Our Complex World. Pegasus Communications. Published online 2001:1–11. Accessed September 18, 2025. www.pegasuscom.com

31. Thompson CJC. The Unintended Consequences of “Having an Impact”. *Syst Thinker*. Published online 2018. Accessed September 18, 2025. https://thesystemsthinker.com/the-unintended-consequences-of-having-an-impact/

32. Schoemaker CG, Richards DP, De Wit M. Matching researchers’ needs and patients’ contributions: Practical tips for meaningful patient engagement from the field of rheumatology. Ann Rheum Dis. 2023;82(3):312–315. doi:10.1136/ard-2022-223561

33. For Patients By Patients. PxP 2025. 2025. Accessed September 23, 2025. https://pxphub.org/event/

34. Meyer E. The Culture Map: Breaking Through the Invisible Boundaries of Global Business. PublicAffairs; 2014. Accessed September 18, 2025. https://books.google.ca/books?id=IMsiBQAAQBAJ&source=gbs_ViewAPI&redir_esc=y

35. U.S. Department of Health and Human Services. Think Cultural Health: Communication Styles.

36. Wu J, Rajesh A, Huang YN, et al. Virtual meetings promise to eliminate geographical and administrative barriers and increase accessibility, diversity and inclusivity. Nature Biotechnology 2021 40:1. 2021;40(1):133-137. doi:10.1038/s41587-021-01176-z

37. Botelho FHF. Accessibility to digital technology: Virtual barriers, real opportunities. Assistive Technology. 2021;33(sup1):27–34. doi:10.1080/10400435.2021.1945705

38. Tully S. A Human Right to Access the Internet? Problems and Prospects. Human Rights Law Review. 2014;14(2):175–195. doi:10.1093/HRLR/NGU011

