## Supplemental File 1. Interview Guide for "Investigating a patient-led conference: What are the characteristics and impacts of patient leadership? A qualitative study"

**SUPPLEMENTAL FILE 2: Interview guide for interviews with SC members**

---

**START OF INTERVIEW**

---

To get started, we'd like to get to know you a bit better through three short questions:

a) Can you please tell us your preferred name and, if you're comfortable, your pronouns?
b) What part of the World you're Zooming in from?
c) And a brief overview of your patient engagement experiences?

1. In your own words, how would you define something that is patient-led?

*Probes: If there was a recipe for patient-led events, what ingredients would be necessary?*

2. Can you tell us about your experience on the PxP steering committee?

*Probes:*

- *How did you contribute? What were your contributions?*
- *Can you describe/talk about your relationship with members of the steering committee? With CIHR/IMHA staff?*
- *Describe the team environment/culture you worked in. Were your ideas implemented?*
- *Can you walk me through the decision-making process for the committee? For example, if someone put forth an idea about a speaker, what happened next? How was the decision made to finalize an idea?*
- *What did meetings look like?*
- *What was it like working in a virtual environment?*
- *What were the strengths of the steering committee?*

3. Do you think PxP was patient-led?

4. During PxP, how were you supported to fulfill your role as a steering committee member?

*Probes*: *What did support look like?*

- *Scheduling of meetings*
- *Accessibility supports (e.g., Closed captioning)*
- *Monetary support (Compensation)*
- *Flexible Meeting times*

- *Technical support (e.g. Zoom walk through)*
- *Access to Zoom platform*

5. What supports were helpful/ not helpful?

*Probe: What supports were missing?*

6. Did you experience any barriers in your role as a steering committee member for PxP?

   6a. <u>If yes,</u> If so, what kinds?

   *Probes:*
   - *Financial? Physical? Emotional?*
   - *Time zones*
   - *Different cultures*
   - *Expectations*
   - *Time commitment*
   - *Energy levels*
   - *Dealing with own illness, life circumstances*

   6b**:** What kind of support would have been helpful to reduce these barriers?

7. How would you compare your PxP steering committee experience to <u>other patient engagement</u> experiences?

8. You defined patient-led at the beginning of the interview – Can you tell us about what you consider different about patient-led initiatives versus other patient engagement opportunities?

*Probes:*
- *Similarities*
- *Differences*
- *Decision making process and power (high, low, same?)*
- *Leadership*
- *Time commitment*
- *Satisfaction or reward*
- *Compensation*
- *Level of Influence / Power dynamics*
- *Level of involvement*
- *Working with different people*

9. What do you think is the impact of patient-led events?

*Probes*:

- *Research community*
- *Impact to themselves*
- *Outside research*
- *Good/Bad*
- *Future Opportunities*
- *Leadership*

10. What role do you think institutions, like IMHA, play in supporting patient-led events?

*Probes*:

- *How did the Institute support?*
- *Funding*
- *Coordination/Secretariat support*
- *Relationship building/connections*
- *Value*
- *Other events the Institute can support*

---

### CLOSING OF INTERVIEW

---

11. Any advice for people involved in future PxP conferences?

12. Is there anything you would like to add before we end today? Or anything you were hoping to discuss that wasn't discussed today?

[STOP RECORDING]
